# Association Patterns of Antisocial Personality Disorder across Substance Use Disorders

**DOI:** 10.1101/2023.09.15.23295625

**Authors:** Aislinn Low, Brendan Stiltner, Yaira Z. Nunez, Keyrun Adhikari, Joseph D. Deak, Robert H. Pietrzak, Henry R. Kranzler, Joel Gelernter, Renato Polimanti

**Affiliations:** Department of Psychiatry, Yale University School of Medicine, New Haven, CT, USA; VA Connecticut Healthcare System, West Haven, CT, USA; U.S. Department of Veterans Affairs National Center for Posttraumatic Stress Disorder, VA Connecticut Healthcare System, West Haven, CT, USA; Department of Psychiatry, University of Pennsylvania Perelman School of Medicine, Philadelphia, PA, USA; Mental Illness Research, Education, and Clinical Center, Crescenz Veterans Affairs Medical Center, Philadelphia, PA, USA; Wu Tsai Institute, Yale University, New Haven, CT, USA

## Abstract

There is a high prevalence of antisocial personality disorder (ASPD) in individuals affected by substance use disorders (SUD). However, there is limited information on the specific patterns of association of ASPD with SUD severity and specific SUD diagnostic criteria. We investigated the association of alcohol, cannabis, cocaine, opioid, and tobacco use disorders (AUD, CanUD, CocUD, OUD, and TUD, respectively) in 1,660 individuals with ASPD and 6,640 controls matched by sex (24% female), age, and racial/ethnic background in a sample ascertained for addiction-related traits. Generalized linear regressions were used to test the association of ASPD with the five DSM-5 SUD diagnoses, their severity (i.e., mild, moderate, severe), and their individual diagnostic criteria. We found that ASPD is associated with the diagnosis and severity of AUD (Odds Ratio, ORs=1.89 and 1.25), CanUD (ORs=2.13 and 1.32), and TUD (ORs=1.50 and 1.21) (*ps*<.003). Of the specific diagnostic criteria, the “hazardous use” criterion showed the strongest association with ASPD across the five SUDs investigated (from OR_TUD_=1.88 to OR_CanUD_=1.37). However, when criteria of different SUDs were included in the same model, ASPD was independently associated only with TUD “hazardous use” and CocUD “attempts to quit”. Attempting to quit cocaine was inversely related to the presence of ASPD and remained significant (OR=0.57, 95% confidence interval = 0.36-0.89) after controlling for interactive effects with sex. The current work provides novel insights into how different SUDs, their severity, and their diagnostic criteria associate with ASPD, potentially furthering our understanding of the impact of polysubstance addiction on mental health.

## INTRODUCTION

Antisocial personality disorder (ASPD) is a psychiatric disorder characterized by a tendency to be manipulative, impulsive, irritable, aggressive, and show a lack of remorse [1]. ASPD is highly comorbid with substance use disorders (SUDs) [1–4]. While nationally representative cohorts report prevalence rates of ASPD around 3.6%, several clinical samples of individuals who misuse substances show a prevalence between 23.5% and 81.4% [5–7]. The higher prevalence of ASPD in the presence of substance misuse supports research on the nature of this relationship and its potential implications for our understanding of the onset, severity, and treatment of both ASPD and SUDs.

Research aimed at understanding the relationship between ASPD and various SUDs has used a variety of experimental designs and sampling techniques. Grant and colleagues (2004) investigated the co-occurrence of alcohol and other drug use disorders (10 different combined drug classes) with seven of the 10 DSM-IV personality disorders in the 2001-2002 wave of the National Epidemiologic Survey on Alcohol and Related Conditions (NESARC; *N*=43,093). Participants in this sample with a current (i.e., past 12 month) alcohol use disorder or other SUD diagnosis were 4.8 and 11.8 times more likely to have an ASPD diagnosis, respectively. In line with this evidence, a study of participants seeking treatment from mental health or drug addiction centers in Spain found that ASPD was the only personality disorder measured that was more prevalent in participants with a lifetime diagnosis of any of the SUDs analyzed (alcohol, 76.5%; cocaine, 81.4%; opiates, 45.1%; cannabis, 67.6%; and sedatives, 23.5%) [6]. Lastly, a study of 200 participants from both inpatient and outpatient psychiatric treatment centers showed that an ASPD diagnosis was associated with a lifetime SUD (a variable that combined hallucinogens, opioids, sedatives, and polysubstance use) even after adjusting for other personality disorders [8].

Building on previous research on the comorbidity between ASPD and various SUD diagnoses, the current study expands our understanding of the complex ASPD-SUD relationship by analyzing the association of ASPD with individual and co-occurring SUDs, testing DSM-5 diagnoses, severity, and diagnostic criteria. Analyses were conducted in the Yale-Penn sample, which, because of its large sample size, deep phenotypic assessment, and purposeful recruitment for SUDs and controls resulting in enrichment for SUD cases, permitted us to investigate the comorbidity of ASPD across the SUD spectrum.

## METHODS

### Yale-Penn Cohort

The Yale-Penn cohort comprises participants recruited to investigate the molecular basis of substance use disorders and other comorbid conditions [9–13]. Yale-Penn participants completed the Semi-Structured Assessment for Drug Dependence and Alcoholism (i.e., SSADDA), a psychiatric research interview administered by trained lay interviewers [14, 15]. The SSADDA is composed of twenty-three sections that assess diagnostic criteria from which diagnoses of SUDs and ASPD can be made.

We investigated DSM-5 diagnoses and individual diagnostic criteria of alcohol use disorder (AUD), cannabis use disorder (CanUD), cocaine use disorder (CocUD), opioid use disorder (OUD), and tobacco use disorder (TUD). We chose to investigate DSM-5 SUDs because it enabled us to analyze the full spectrum of addiction-related behaviors without distinguishing between substance abuse and dependence as defined by DSM-IV. DSM-5 SUD criteria included: a) hazardous use, b) social problems due to use, c) neglected roles due to use, d) withdrawal, e) tolerance, f) using larger amounts, g) attempts to quit, h) much time spent using, i) physical problems due to use, j) activities given up due to use, and k) craving [1]. The severity of each SUD was estimated by summing the respective diagnostic criteria (coded as 1 = present and 0 = not present) and classifying each participant based on the DSM-5 definition for mild (i.e., 2-3 criteria present), moderate (i.e., 4-5 criteria present), and severe (i.e., 6 or more criteria present) SUD. Because the SSADDA was originally designed to collect information related to DSM-IV criteria for substance abuse and dependence, which did not include tobacco abuse, we could not derive the DSM-5 TUD criteria b and c. In contrast, we analyzed DSM-IV diagnoses for ASPD (diagnosed independent of substance-induced behaviors), major depressive disorder (MDD), generalized anxiety disorder (GAD), and posttraumatic stress disorder (PTSD) because the SSADDA, which was designed to assess DSM-IV diagnoses, does not allow translation to DSM-5 for these disorders. are only minor differences between DSM-IV and DSM-5 criteria for these mental illnesses. We also extracted data regarding sex, age, race/ethnicity, education, and household income.

### Case-Control Matching

A total of 1,660 Yale-Penn participants received a lifetime ASPD diagnosis. To maximize the statistical power of the analysis and to control for the effect of demographic characteristics on ASPD-SUD associations, we matched ASPD cases with controls with no ASPD diagnosis on sex, age, and race/ethnicity. ASPD case-control matching was performed in R Statistical Software (v4.2.2) using the MatchIt package (v4.5.2) [16]. We estimated only a 2%-increase in the effective sample size after reaching a 1:7 case-control ratio (Supplemental Table 1). Chi-square and t-test analyses were performed where appropriate on the matching criteria to confirm acceptable matching in the sample. Appropriate matching was observed at the 1:4 case-control ratio, which corresponded to 1,660 ASPD cases and 6,640 controls (Table 1).

**Table 1:** Characteristics of ASPD cases and controls from Yale-Penn cohort. Controls were matched to cases for sex, age, and race-ethnicity.

| Characteristics | Cases<br>(*N* = 1,660) | Controls<br>(*N* = 6,640) | *p* |
| --- | --- | --- | --- |
| Sex, *N* (%) |  |  |  |
| Male | 1,256 (76) | 5,044 (76) | .822 |
| Female | 404 (24) | 1,596 (24) |  |
| Age |  |  |  |
| Years (SD) | 38.23 (11) | 38.30 (11) | .822 |
| Race-Ethnicity, *N* (%) |  |  |  |
| Native American | 11 (1) | 40 (1) | .609 |
| Asian | 3 (<1) | 10 (<1) |  |
| Pacific Islander | 4 (<1) | 11 (<1) |  |
| African American, no Hispanic descent | 681 (41) | 2,859 (43) |  |
| African American, Hispanic descent | 54 (3) | 185 (3) |  |
| European-American, no Hispanic descent | 657 (40) | 2,641 (40) |  |
| European-American, Hispanic descent | 118 (7) | 413 (6) |  |
| Other | 132 (8) | 481 (7) |  |
| Education Level, *N* (%) |  |  |  |
| Less than High School | 130 (8) | 198 (3) | 1.33 × 10<sup>-68</sup> |
| High School | 1,090 (66) | 3,615 (54) |  |
| Some College | 357 (22) | 1,782 (27) |  |
| Bachelor's or Higher | 83 (4) | 1,045 (16) |  |
| Annual Household Income, *N* (%) |  |  |  |
| \$0 - \$9,999 | 890 (54) | 2,974 (45) | 4.06 × 10<sup>-16</sup> |
| \$10,000 - \$19,999 | 320 (19) | 1,163 (18) | |
| \$20,000 - \$29,999 | 169 (10) | 776 (12) | |
| \$30,000 - \$39,999 | 109 (7) | 497 (7) | |
| \$40,000 - \$49,999 | 67 (4) | 326 (5) | |
| \$50,000 - \$74,999 | 59 (4) | 436 (6) | |
| \$75,000 - \$99,999 | 20 (1) | 208 (3) | |
| \$100,000 - \$149,999 | 18 (1) | 178 (3) | |
| \$150,000+ | 8 (<1) | 82 (1) | |

### Statistical Analysis

The association between ASPD and SUDs were tested using generalized linear regression, in which AUD, CanUD, CocUD, OUD, and TUD were considered in terms of diagnosis (case-control status), severity (mild, moderate, and severe), and individual diagnostic criteria. We included education, household income, age, sex, and race-ethnicity as covariates to account for residual effects of demographic characteristics. To control for the comorbidity of other common mental health disorders, we also included internalizing disorders (i.e., MDD, GAD, and PTSD) in the regression model. Variables related to different SUDs (i.e., AUD, CanUD, CocUD, OUD, and TUD) were also tested in a single omnibus model to account for polysubstance addiction. Bonferroni correction was applied to account for the multiple association tests performed.

## RESULTS

### Characteristics of ASPD Cases and Controls

As expected, based on matching on these characteristics, there were no statistically significant differences between ASPD cases and controls on sex, age, or race-ethnicity (*p* > 0.6; Table 1). The overall sample (*N* = 8,300) overrepresented males (>75%) to reflect the epidemiology of ASPD [17]. There were comparable proportions of African Americans (43%) and European Americans (40%), with Hispanics comprising <10% of the sample. ASPD cases reported lower household income and education level than controls (*ps* = 4.06_X_10^-16^ and 1.33_X_10^-68^, respectively; Table 1). Overall, most of the sample had an annual household income ≤$9,999 (47%) and reported high school as the highest degree completed (57%).

### ASPD Association with SUD Diagnoses and Severity Measures

Most participants (*N* = 8,300; Supplemental Table 2) had a lifetime diagnosis of AUD (72%), TUD (70%), CocUD (64%), CanUD (51%), or OUD (39%). Controlling for sex, age, race-ethnicity, income, education, and the five SUDs, we observed ASPD-SUD associations that survived Bonferroni correction (*p* < .003; Supplemental Table 3) for AUD (OR=1.94, 95%CI=1.63-2.31), CanUD (OR=2.13, 95%CI=1.94-2.52), and TUD (OR=1.53, 95%CI=1.28-1.82). Among the covariates included in the model, sex was associated with ASPD (*p* = 1.60_X_10^-5^; Supplemental Table 3) despite the ASPD cases and controls being well matched on sex (Table 1). This suggests that there are sex differences related to the co-occurrence of ASPD and SUD. Thus, we included GAD, MDD, and PTSD diagnoses as additional covariates, which although not changing the ASPD-SUD associations, attenuated the association with sex (Supplemental Table 3). We chose to include internalizing disorders as covariates in the subsequent analyses to control for possible residual confounding factors. Testing the effect of SUD severity on the association with ASPD demonstrated a pattern similar to that found for the diagnosis-based analysis (Figure 1; Supplemental Table 4). However, SUD diagnoses were more strongly associated with ASPD than SUD severity measures (diagnosis vs. severity Δ*p* ≤ 1.54_X_10^-4^).

**Fig. 1:**
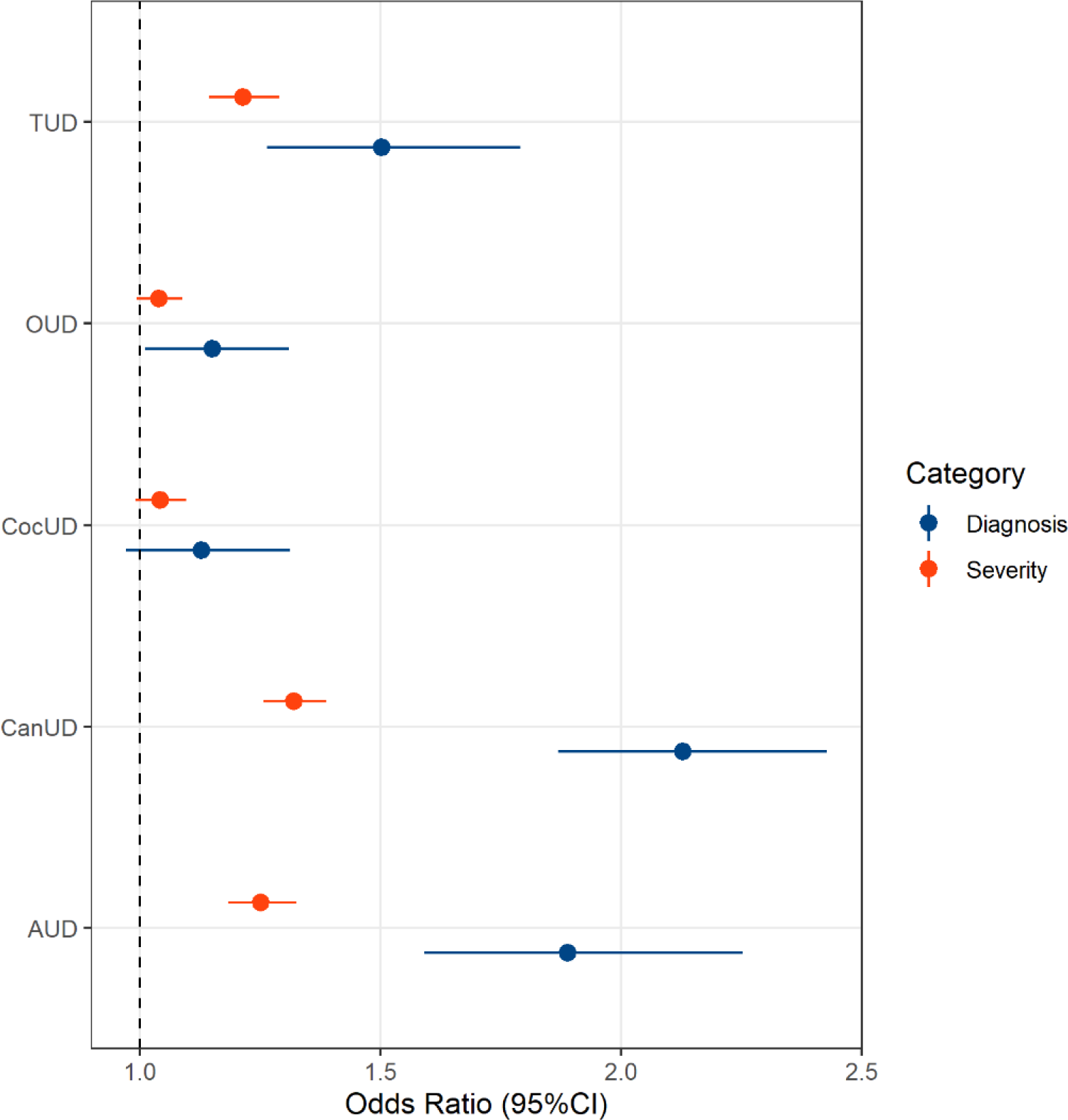
Association of antisocial personality disorder with alcohol use disorder (AUD), cannabis use disorder (CanUD), cocaine use disorder (CocUD), opioid use disorder (OUD), and tobacco use disorder (TUD).

### ASPD Associations with SUD Diagnostic Criteria

To test whether certain SUD features are associated with ASPD, we investigated individual diagnostic criteria for each of the five SUDs of interest. These models included SUD severity, demographic characteristics, socioeconomic factors, and internalizing disorders as covariates. Analyzing each SUD separately, we identified associations between ASPD and 12 substance-specific diagnostic criterion that survived Bonferroni correction (*p* < .002; Figure 2; Supplemental Tables 5-9). The “Hazardous Use” criterion was statistically significant across all SUDs investigated (OR_AUD_ = 1.83, OR_CanUD_ = 1.37, OR_CocUD_ = 1.63, OR_OUD_ = 1.73, and OR_TUD_ = 1.88, *ps* < .002). Significant associations were seen for the “Social Problems” criterion in both AUD (OR = 1.55, *p* = 4.99_X_10^-5^) and CanUD (OR = 1.28, *p* = 1.67_X_10^-3^), the “Neglected Roles” criterion in both CanUD (OR = 1.31, *p* = 1.86_X_10^-3^) and CocUD (OR = 1.35, *p* = 1.62_X_10^-3^), the “Attempts to Quit” criterion in both AUD (OR = .76, *p* = 9.30_X_10^-4^) and CocUD (OR = .57, *p* = 1.48_X_10^-5^), and the “Physical Problems” criterion in TUD (OR = 1.29, *p* = 6.16_X_10^-4^).

**Fig. 2:**
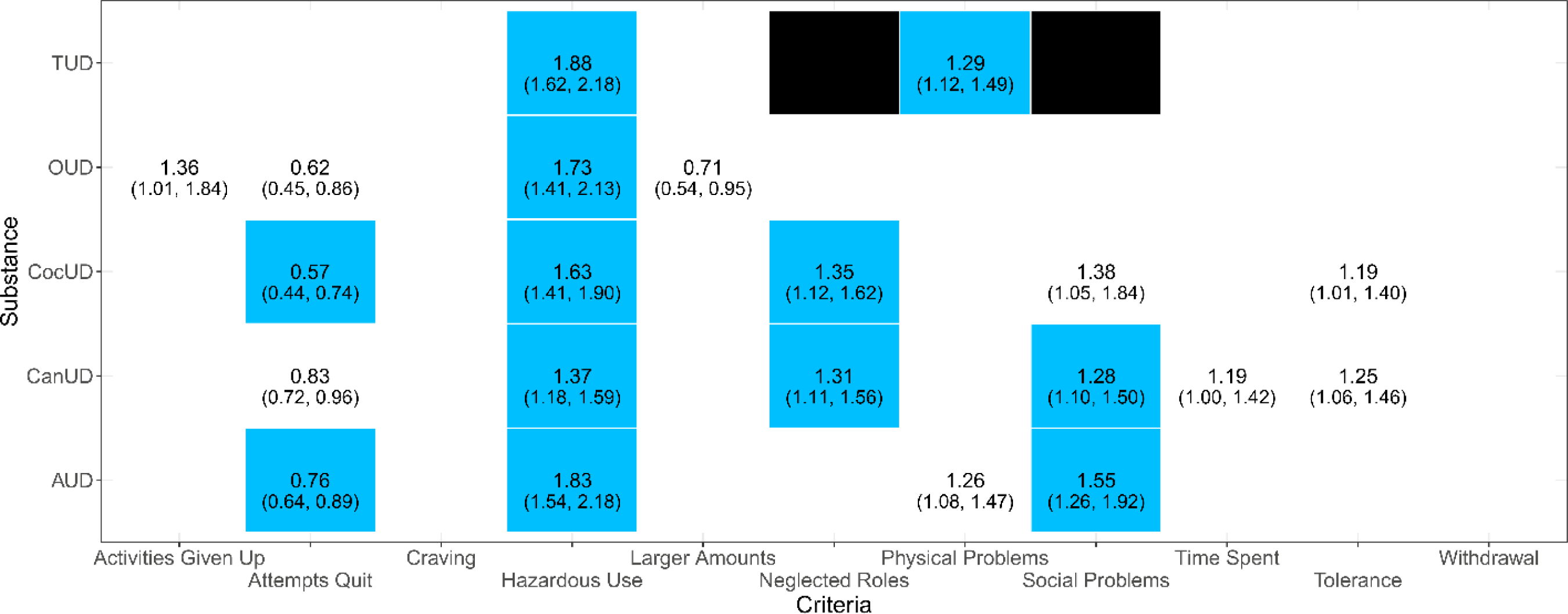
Associations of antisocial personality disorder with individual diagnostic criteria of alcohol use disorder (AUD), cannabis use disorder (CanUD), cocaine use disorder (CocUD), opioid use disorder (OUD), and tobacco use disorder (TUD). Blue cells represent associations surviving Bonferroni correction (*p* < .002). White cells with odds ratio statistics represent nominally significant associations (p<0.05). Empty cells represent null associations (p>0.05). Black cells represent TUD diagnostic criteria not available.

To account for comorbidity across SUDs, we reanalyzed the significant diagnostic criteria in each SUD-specific model, entering them in a regression model together with the previously listed covariates (Supplemental Table 10). In this multi-SUD analysis, three SUD-related phenotypes survived Bonferroni correction (*p* < 1.61_X_10^-3^; Figure 3): CanUD severity (OR = 1.17, *p* = 3.66_X_10^-5^), CocUD “Attempts to Quit” (OR = .64, *p* = 8_X_10^-4^), and TUD “Hazardous Use” (OR = 1.54, *p* = 7.38_X_10^-9^). However, among the covariates included in the model, sex was associated with ASPD (*p* = 1.15_X_10^-4^; Supplemental Table 11) despite being matched between ASPD cases and controls (Table 1). To account for this residual effect of sex, we entered sex interaction terms in the model for the three SUD-related phenotypes (Supplemental Table 11). After accounting for sex interactions, only CocUD “Attempts to Quit” was still negatively associated with ASPD (OR = .57, 95%CI = 0.36-0.89, *p* = .013; Figure 3).

**Fig. 3:**
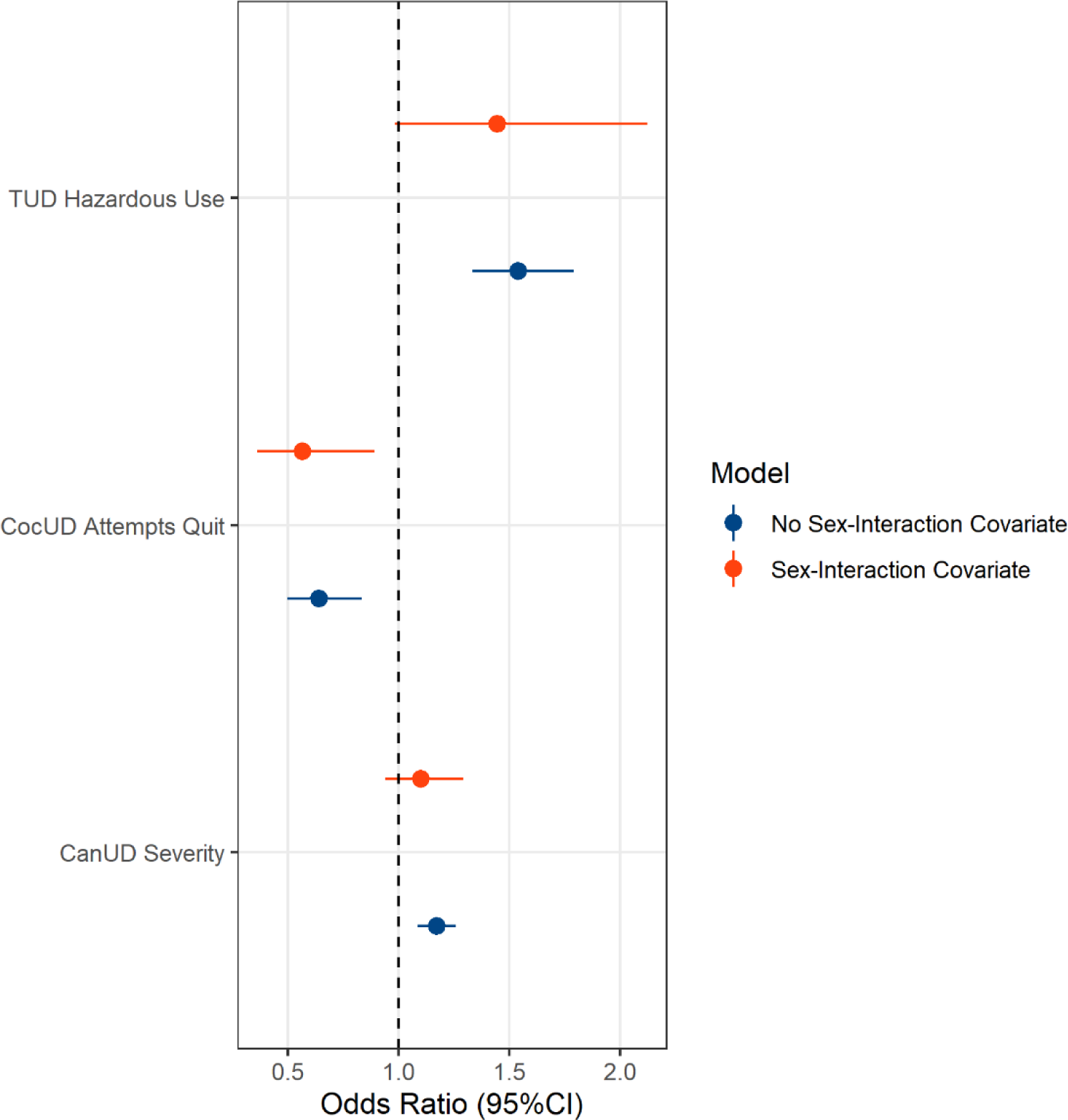
Associations of antisocial personality disorder with individual diagnostic criteria across multiple substance use disorders. Criteria surviving Bonferroni multiple testing correction are reported before and after including sex-interaction covariates. The results of the full models are reported in Supplemental Tables 10 and 11.

## DISCUSSION

We investigated associations of ASPD with a broad spectrum of SUD related outcomes. We did this by modeling ASPD in relation to SUD diagnostic status, both independently and simultaneously (i.e., all SUDs included in the same model), SUD severity thresholds, and individual substance-specific diagnostic criteria.

We found that diagnoses and severity measures of AUD, CanUD, CocUD, OUD, and TUD were positively associated with an ASPD diagnosis, although only the associations with AUD, CanUD, and TUD survived multiple testing correction. This may be due to the higher population prevalence of those three diagnoses than for CocUD and OUD [18]. The diagnosis-based associations are consistent with previous findings of SUDs being positively associated with ASPD [19, 20]. This association may arise from shared etiologies and predispositions between ASPD and SUDs, as well as other externalizing disorders [21]. One common feature linked to both ASPD and SUDs is behavioral disinhibition [21–23], which is loosely composed of sensation seeking, impulsivity, and aggressivity [24, 25]. AUD specifically has been linked to low harm avoidance, high novelty seeking, and low reward dependence, which are characteristics shared with ASPD [26–28]. It is reasonable to infer that these behaviors may extend beyond alcohol-related traits to include other substance-related traits consistent with our findings that ASPD is associated with drug use disorder diagnoses and measures of severity.

This is the first investigation of which we are aware that assesses the specific associations of ASPD with the individual diagnostic criteria used to diagnose SUDs. The criteria that survived multiple testing correction in the SUD-specific analyses (“Hazardous Use”, “Social Problems”, “Neglected Roles”, “Physical Problems”, and “Attempts to Quit”; Figure 2) align well with the impulsive, reckless, and irresponsible characteristics of ASPD. “Hazardous Use” has been linked to measures of disinhibition and antisocial behaviors [29, 30] and is the only criterion to associate across all the SUDs, indicating that individuals with co-occurring ASPD and SUD use substances in hazardous situations, regardless of the substance. This is consistent with the conceptualization of the “Hazardous Use” criterion as measuring general externalizing behavior independent of the substance itself and varying between demographic groups [31, 32]. Notably, the “Attempts to Quit” criterion was the only one that was negatively associated with ASPD, suggesting that the absence of ASPD, or decreased ASPD symptomology, allows individuals to make more attempts to terminate their substance use. However, there are different views on whether the presence of ASPD supports or hinders SUD treatment seeking behavior and abstinence [33–35]. Further, the different associations of ASPD with criteria for individual SUDs supports our decision to analyze each separately rather than creating a composite SUD variable for analysis.

While significant when tested individually, most associations related to SUD criteria were null when combined into a multi-SUD analysis of diagnostic criteria. This is likely due to the intercorrelation among diagnostic criteria across SUDs [36]. Sex was significantly associated with ASPD/SUD associations despite having controlled for sex differences in ASPD prevalence by matching cases and controls. The association with sex was attenuated when internalizing disorders were included as covariates in the regression analysis. ASPD [17, 37], SUDs [38, 39], and internalizing disorders (i.e., MDD, GAD, and PTSD) [40–42], and their respective comorbidities [43, 44] all have distinct sex distributions. Despite controlling for sex across ASPD cases and controls, the re-emergence of sex as a significant covariate in the regression analysis of multi-SUD, diagnostic criteria regression likely arose from symptom-level sex differences in SUD criteria that exceed those at the SUD diagnostic or severity level. The inclusion of sex-interaction terms in the analysis of significant SUD criteria (e.g., Sex-TUD Hazardous Use, Sex-CocUD Attempts to Quit, and Sex-CanUD Severity) controlled for these sex effects.

This study has several limitations. Despite providing a large, racially diverse sample of participants, recruitment of the Yale-Penn cohort targeted individuals with addiction phenotypes (and controls without these phenotypes). Thus, results from these analyses cannot be generalized to the general population or other cohorts with differing characteristics. However, they bring to light specific ASPD associations across the SUD spectrum and underscore the potential utility of conducting similar analyses in a nationally representative cohort for comparison. Secondly, although the goal of the present study was to identify ASPD associations across the SUD spectrum, the current design does not permit us to investigate the cause-effect relationships that may contribute to the associations observed. Thirdly, only DSM-IV criteria were available for ASPD, MDD, GAD, and PTSD. We do not anticipate any significant impact on the analyses from this as there were only minor differences between DSM-IV and DSM-5 for these disorders. Lastly, two DSM-5 criteria for TUD (i.e., “Social Problems” and “Neglected Roles”) are not available in the SSADDA data. Excluding these criteria could have led to an under-identification of individuals with TUD and misclassification of TUD severity. Additionally, it may directly impact the TUD-specific and SUD-combined criteria analyses as “Social Problems” and “Neglected Roles” were seen to be significantly associated with ASPD in multiple SUDs.

Future studies will be needed to replicate these findings in cohorts selected for characteristics other than substance use phenotypes, which would show how generalizable the current findings are. It may also be helpful to assess whether similar associations exist between SUDs and other externalizing disorders that comprise ASPD: namely conduct disorder [45] and adult antisocial behavior [46, 47]. These analyses could enhance our understanding of the extent to which these associations reflect overall deviance from cultural norms and mental health comorbidities of the population at large.

In conclusion, the comorbidity of ASPD and SUD is of significant theoretical and clinical interest. This study builds on previous work in this area by showing consistent associations between ASPD and SUD diagnoses and uncovering unique, substance-specific associations between ASPD and individual SUD diagnostic criteria. The present study expands our understanding of polysubstance addiction and co-occurring mental health disorders. Future work in other samples, particularly general population samples, is needed to confirm and extend these associations.

## Data Availability

All data produced in the present work are contained in the manuscript.

## ACKNOWLEDGMENTS

The authors thank the research participants enrolled in the Yale-Penn cohort. This study was supported by the National Institutes of Health (R33 DA047527, R21 DC018098, RF1 MH132337, and T32 AA028259), One Mind, and the VISN 4 Mental Illness Research, Education and Clinical Center at the Crescenz VAMC. The Yale-Penn cohort was supported by multiple grants from the National Institutes of Health (RC2 DA028909, R01 DA12690, R01 DA12849, R01 DA18432, R01 AA11330, R01 AA017535). The funding sources had no role in the design of this study, its execution, analyses, interpretation of the data, and the decision to publish the results.

