## Supplemental Tables for "Association Patterns of Antisocial Personality Disorder across Substance Use Disorders"

**Supplemental Table 1**: Increase in effective sample size considering different case-control ratios. The % increase is calculated with respect to the antecedent case-control ratio.

| **case:control ratio** | **cases** | **controls** | **effective sample size** | **%increase** |
| --- | --- | --- | --- | --- |
| 1:1 | 1660 | 1660 | 3320 | - |
| 1:2 | 1660 | 3320 | 4427 | 33% |
| 1:3 | 1660 | 4980 | 4980 | 13% |
| 1:4 | 1660 | 6640 | 5312 | 7% |
| 1:5 | 1660 | 8300 | 5533 | 4% |
| 1:6 | 1660 | 9960 | 5691 | 3% |
| 1:7 | 1660 | 11620 | 5810 | 2% |
| 1:8 | 1660 | 13280 | 5902 | 2% |
| 1:9 | 1660 | 14940 | 5976 | 1% |
| 1:10 | 1660 | 16600 | 6036 | 1% |

**Supplemental Table 2**: Distribution of substance use disorders and internalizing disorders between ASPD cases and controls. AUD: alcohol use disorder; CanUD: cannabis use disorder; CocUD: cocaine use disorder; OUD: opioid use disorder; TUD: tobacco use disorder; MDD: major depressive disorder; GAD: generalized anxiety disorder; PTSD: posttraumatic stress disorder.

| **Diagnosis** N (%) | **Cases** (N=1,660) | **Controls** (N=6,640) |
| --- | --- | --- |
| **AUD** | **1,464 (88)** | **4,526 (68)** |
| *Mild* | *137 (8)* | *819 (12)* |
| *Moderate* | *181 (11)* | *730 (11)* |
| *Severe* | *1,146 (69)* | *2,977 (45)* |
| **CanUD** | **1,202 (72)** | **2,992 (45)** |
| *Mild* | *254 (15)* | *930 (14)* |
| *Moderate* | *264 (16)* | *739 (11)* |
| *Severe* | *684 (41)* | *1,323 (20)* |
| **CocUD** | **1,301 (78)** | **3,995 (60)** |
| *Mild* | *41 (2)* | *274 (4)* |
| *Moderate* | *85 (5)* | *368 (6)* |
| *Severe* | *1,175 (71)* | *3,353 (51)* |
| **OUD** | **836 (50)** | **2,364 (36)** |
| *Mild* | *40 (2)* | *146 (2)* |
| *Moderate* | *44 (3)* | *171 (3)* |
| *Severe* | *752 (45)* | *2,047 (31)* |
| **TUD** | **1,444 (87)** | **4,380 (66)** |
| *Mild* | *142 (9)* | *875 (13)* |
| *Moderate* | *364 (22)* | *1,375 (21)* |
| *Severe* | *938 (57)* | *2,130 (32)* |
| **MDD** | **335 (20)** | **760 (11)** |
| **GAD** | **22 (1)** | **65 (1)** |
| **PTSD** | **348 (21)** | **729 (11)** |

**Supplemental Table 3:** Association of antisocial personality disorder with alcohol use disorder (AUD), cannabis use disorder (CanUD), cocaine use disorder (CocUD), opioid use disorder (OUD), and tobacco use disorder (TUD). Associations surviving Bonferroni multiple testing correction (p < .003) are indicated in red. Covariates and phenotypes of interest are highlighted in orange and blue, respectively. NOHO: Not of Hispanic Origin; OHO: Of Hispanic Origin; MDD: major depressive disorder; GAD: generalized anxiety disorder; PTSD: posttraumatic stress disorder.

| **Variables** | **Model 1** | | | | **Model 2** | | | |
| --- | --- | --- | --- | --- | --- | --- | --- | --- |
|  | **OR** | **LCI** | **UCI** | ***p*** | **OR** | **LCI** | **UCI** | ***p*** |
| Sex | **1.35** | **1.18** | **1.55** | **1.60E-05** | 1.18 | 1.03 | 1.36 | 0.018 |
| Age | 1.00 | 1.00 | 1.01 | 0.384 | 1.00 | 1.00 | 1.01 | 0.579 |
| Native American | 1.11 | 0.53 | 2.31 | 0.787 | 1.23 | 0.57 | 2.49 | 0.584 |
| Asian | 4.33 | 1.02 | 18.48 | 0.048 | 3.57 | 0.65 | 15.29 | 0.108 |
| Pacific Islander | 2.14 | 0.61 | 7.52 | 0.234 | 2.63 | 0.67 | 8.61 | 0.128 |
| African American, NOHO | 0.96 | 0.77 | 1.20 | 0.745 | 1.05 | 0.84 | 1.32 | 0.680 |
| African American, OHO | 1.00 | 0.69 | 1.46 | 0.998 | 1.00 | 0.68 | 1.46 | 0.994 |
| European American, NOHO | 1.04 | 0.83 | 1.30 | 0.741 | 1.10 | 0.87 | 1.38 | 0.434 |
| European American, OHO | 0.94 | 0.70 | 1.27 | 0.699 | 0.95 | 0.71 | 1.28 | 0.753 |
| Education Level | **0.85** | **0.83** | **0.88** | **2.30E-25** | **0.85** | **0.83** | **0.88** | **1.36E-25** |
| Household Income | 1.00 | 0.96 | 1.03 | 0.838 | 1.00 | 0.96 | 1.03 | 0.788 |
| MDD | - | - | - | - | **1.72** | **1.47** | **2.01** | **6.62E-12** |
| GAD | - | - | - | - | 1.25 | 0.73 | 2.09 | 0.400 |
| PTSD | - | - | - | - | **1.51** | **1.29** | **1.75** | **1.44E-07** |
| AUD | **1.94** | **1.63** | **2.31** | **6.26E-14** | **1.89** | **1.59** | **2.25** | **7.10E-13** |
| CanUD | **2.12** | **1.94** | **2.52** | **4.92E-33** | **2.13** | **1.87** | **2.43** | **1.12E-29** |
| CocUD | 1.12 | 0.97 | 1.30 | 0.132 | 1.13 | 0.97 | 1.31 | 0.114 |
| OUD | 1.15 | 1.01 | 1.31 | 0.030 | 1.15 | 1.01 | 1.31 | 0.033 |
| TUD | **1.53** | **1.28** | **1.82** | **1.71E-06** | **1.50** | **1.26** | **1.79** | **4.41E-06** |

**Supplemental Table 4.** Association of antisocial personality disorder with severity of alcohol use disorder (AUD), cannabis use disorder (CanUD), cocaine use disorder (CocUD), opioid use disorder (OUD), and tobacco use disorder (TUD). Associations surviving Bonferroni multiple testing correction (p < .003) are indicated in red. Covariates and phenotypes of interest are highlighted in orange and blue, respectively. NOHO: Not of Hispanic Origin; OHO: Of Hispanic Origin; MDD: major depressive disorder; GAD: generalized anxiety disorder; PTSD: posttraumatic stress disorder.

| **Variable** | **OR** | **LCI** | **UCI** | ***p*** |
| --- | --- | --- | --- | --- |
| Sex | 1.21 | 1.05 | 1.39 | 0.008 |
| Age | 1.00 | 1.00 | 1.01 | 0.668 |
| Native American | 1.32 | 0.60 | 2.69 | 0.467 |
| Asian | 3.88 | 0.73 | 16.08 | 0.080 |
| Pacific Islander | 2.74 | 0.70 | 8.99 | 0.112 |
| African American, NOHO | 1.10 | 0.87 | 1.38 | 0.434 |
| African American, OHO | 1.03 | 0.70 | 1.51 | 0.881 |
| European American, NOHO | 1.11 | 0.88 | 1.40 | 0.389 |
| European American, OHO | 0.97 | 0.72 | 1.31 | 0.831 |
| **Education Level** | **0.86** | **0.83** | **0.88** | **5.20E-24** |
| Household Income | 1.00 | 0.97 | 1.04 | 0.835 |
| **MDD** | **1.64** | **1.40** | **1.92** | **5.25E-10** |
| GAD | 1.27 | 0.73 | 2.13 | 0.374 |
| **PTSD** | **1.36** | **1.16** | **1.58** | **1.14E-04** |
| **AUD** | **1.25** | **1.18** | **1.33** | **4.07E-15** |
| **CanUD** | **1.32** | **1.26** | **1.39** | **3.24E-28** |
| CocUD | 1.04 | 0.99 | 1.10 | 0.110 |
| OUD | 1.04 | 0.99 | 1.09 | 0.085 |
| **TUD** | **1.21** | **1.14** | **1.29** | **1.89E-10** |

**Supplemental Table 5.** Association of antisocial personality disorder with severity and diagnostic criteria of alcohol use disorder (AUD). Associations surviving Bonferroni multiple testing correction (p < .002) are indicated in red. Covariates and phenotypes of interest are highlighted in orange and blue, respectively. NOHO: Not of Hispanic Origin; OHO: Of Hispanic Origin; MDD: major depressive disorder; GAD: generalized anxiety disorder; PTSD: posttraumatic stress disorder.

| **Variable** | | | **OR** | **LCI** | **UCI** | | ***p*** |
| --- | --- | --- | --- | --- | --- | --- | --- |
| Sex | | | 1.13 | 0.99 | 1.30 | | 0.079 |
| Age | | | 1.00 | 0.99 | 1.00 | | 0.356 |
| Native American | | | 1.25 | 0.57 | 2.55 | | 0.552 |
| Asian | | | 3.14 | 0.61 | 12.50 | | 0.128 |
| Pacific Islander | | | 2.58 | 0.67 | 8.33 | | 0.131 |
| African American, NOHO | | | 1.08 | 0.86 | 1.35 | | 0.519 |
| African American, OHO | | | 1.11 | 0.75 | 1.62 | | 0.598 |
| European American, NOHO | | | 1.06 | 0.85 | 1.34 | | 0.602 |
| European American, OHO | | | 1.00 | 0.74 | 1.36 | | 0.974 |
| **Education Level** | | | **0.83** | **0.80** | **0.85** | | **3.61E-38** |
| Household Income | | | 0.98 | 0.94 | 1.01 | | 0.169 |
| **MDD** | | | **1.70** | **1.46** | **1.99** | | **2.03E-11** |
| GAD | | | 1.10 | 0.64 | 1.82 | | 0.727 |
| **PTSD** | | | **1.53** | **1.31** | **1.78** | | **7.36E-08** |
| **AUD** | Severity | 0.97 | | 0.83 | 1.15 | 0.761 | |
|  | **Hazardous** | **1.83** | | **1.54** | **2.18** | **1.47E-11** | |
|  | **Social Problems** | **1.55** | | **1.26** | **1.92** | **4.99E-05** | |
|  | Neglected Roles | 1.14 | | 0.95 | 1.36 | 0.161 | |
|  | Withdrawal | 1.09 | | 0.93 | 1.27 | 0.301 | |
|  | Tolerance | 1.15 | | 0.98 | 1.34 | 0.082 | |
|  | Larger Amounts | 1.22 | | 0.98 | 1.53 | 0.078 | |
|  | **Attempts Quit** | **0.76** | | **0.64** | **0.89** | **9.30E-04** | |
|  | Time Spent | 0.94 | | 0.81 | 1.10 | 0.453 | |
|  | Physical Problems | 1.26 | | 1.08 | 1.47 | 0.003 | |
|  | Activities Given Up | 1.05 | | 0.89 | 1.24 | 0.543 | |
|  | Craving | 0.90 | | 0.78 | 1.05 | 0.171 | |

**Supplemental Table 6.** Association of antisocial personality disorder with severity and diagnostic criteria of cannabis use disorder (CanUD). Associations surviving Bonferroni multiple testing correction (p < .002) are indicated in red. Covariates and phenotypes of interest are highlighted in orange and blue, respectively. NOHO: Not of Hispanic Origin; OHO: Of Hispanic Origin; MDD: major depressive disorder; GAD: generalized anxiety disorder; PTSD: posttraumatic stress disorder.

| **Variable** | | | **OR** | | **LCI** | | **UCI** | | ***p*** |
| --- | --- | --- | --- | --- | --- | --- | --- | --- | --- |
| Sex | | | 1.22 | | 1.06 | | 1.41 | | 0.005 |
| Age | | | 1.01 | | 1.00 | | 1.01 | | 0.071 |
| Native American | | | 1.34 | | 0.62 | | 2.69 | | 0.436 |
| Asian | | | 3.40 | | 0.66 | | 13.18 | | 0.100 |
| Pacific Islander | | | 3.01 | | 0.78 | | 9.52 | | 0.077 |
| African American, NOHO | | | 1.07 | | 0.86 | | 1.35 | | 0.551 |
| African American, OHO | | | 1.07 | | 0.72 | | 1.56 | | 0.737 |
| European American, NOHO | | | 1.18 | | 0.94 | | 1.48 | | 0.161 |
| European American, OHO | | | 1.00 | | 0.74 | | 1.35 | | 0.987 |
| **Education Level** | | | **0.82** | | **0.80** | | **0.84** | | **1.29E-41** |
| Household Income | | | 0.97 | | 0.94 | | 1.00 | | 0.065 |
| **MDD** | | | **1.68** | | **1.44** | | **1.97** | | **7.72E-11** |
| GAD | | | 1.15 | | 0.66 | | 1.93 | | 0.602 |
| **PTSD** | | | **1.46** | | **1.25** | | **1.71** | | **1.74E-06** |
| **CanUD** | Severity | 1.17 | | 1.00 | | 1.35 | | 0.045 | |
|  | **Hazardous** | **1.37** | | **1.18** | | **1.59** | | **3.80E-05** | |
|  | **Social Problems** | **1.28** | | **1.10** | | **1.50** | | **1.67E-03** | |
|  | **Neglected Roles** | **1.31** | | **1.11** | | **1.56** | | **1.86E-03** | |
|  | Withdrawal | 0.96 | | 0.81 | | 1.13 | | 0.619 | |
|  | Tolerance | 1.25 | | 1.06 | | 1.46 | | 0.007 | |
|  | Larger Amounts | 1.00 | | 0.84 | | 1.18 | | 0.967 | |
|  | Attempts Quit | 0.83 | | 0.72 | | 0.96 | | 0.014 | |
|  | Time Spent | 1.19 | | 1.00 | | 1.42 | | 0.046 | |
|  | Physical Problems | 1.13 | | 0.96 | | 1.33 | | 0.136 | |
|  | Activities Given Up | 0.96 | | 0.81 | | 1.14 | | 0.671 | |
|  | Craving | 1.00 | | 0.85 | | 1.17 | | 0.997 | |

**Supplemental Table 7.** Association of antisocial personality disorder with severity and diagnostic criteria of cocaine use disorder (CocUD). Associations surviving Bonferroni multiple testing correction (p < .002) are indicated in red. Covariates and phenotypes of interest are highlighted in orange and blue, respectively. NOHO: Not of Hispanic Origin; OHO: Of Hispanic Origin; MDD: major depressive disorder; GAD: generalized anxiety disorder; PTSD: posttraumatic stress disorder.

| **Variable** | | **OR** | **LCI** | **UCI** | ***p*** |
| --- | --- | --- | --- | --- | --- |
| Sex | | 1.01 | 0.88 | 1.16 | 0.846 |
| Age | | 0.99 | 0.99 | 1.00 | 0.016 |
| Native American | | 1.44 | 0.67 | 2.91 | 0.324 |
| Asian | | 2.48 | 0.47 | 9.77 | 0.229 |
| Pacific Islander | | 2.69 | 0.71 | 8.36 | 0.107 |
| African American, NOHO | | 1.10 | 0.88 | 1.38 | 0.395 |
| African American, OHO | | 1.08 | 0.74 | 1.58 | 0.674 |
| European American, NOHO | | 1.19 | 0.95 | 1.49 | 0.137 |
| European American, OHO | | 0.98 | 0.73 | 1.32 | 0.890 |
| **Education Level** | | **0.82** | **0.80** | **0.84** | **4.32E-41** |
| Household Income | | 0.96 | 0.93 | 0.99 | 0.018 |
| **MDD** | | **1.75** | **1.50** | **2.04** | **1.05E-12** |
| GAD | | 1.18 | 0.69 | 1.96 | 0.527 |
| **PTSD** | | **1.54** | **1.32** | **1.79** | **3.49E-08** |
| **CocUD** | Severity | 0.93 | 0.77 | 1.12 | 0.473 |
|  | **Hazardous** | **1.63** | **1.41** | **1.90** | **2.18E-10** |
|  | Social Problems | 1.38 | 1.05 | 1.84 | 0.024 |
|  | **Neglected Roles** | **1.35** | **1.12** | **1.62** | **1.62E-03** |
|  | Withdrawal | 1.15 | 0.95 | 1.39 | 0.165 |
|  | Tolerance | 1.19 | 1.01 | 1.40 | 0.035 |
|  | Larger Amounts | 1.09 | 0.85 | 1.39 | 0.503 |
|  | **Attempts Quit** | **0.57** | **0.44** | **0.74** | **1.48E-05** |
|  | Time Spent | 0.91 | 0.74 | 1.14 | 0.413 |
|  | Physical Problems | 1.13 | 0.84 | 1.53 | 0.415 |
|  | Activities Given Up | 0.98 | 0.79 | 1.21 | 0.834 |
|  | Craving | 1.13 | 0.94 | 1.35 | 0.191 |

**Supplemental Table 8.** Association of antisocial personality disorder with severity and diagnostic criteria of opioid use disorder (OUD). Associations surviving Bonferroni multiple testing correction (p < .002) are indicated in red. Covariates and phenotypes of interest are highlighted in orange and blue, respectively. NOHO: Not of Hispanic Origin; OHO: Of Hispanic Origin; MDD: major depressive disorder; GAD: generalized anxiety disorder; PTSD: posttraumatic stress disorder.

| **Variable** | | **OR** | **LCI** | **UCI** | ***p*** |
| --- | --- | --- | --- | --- | --- |
| Sex | | 0.98 | 0.85 | 1.12 | 0.762 |
| Age | | 1.00 | 0.99 | 1.00 | 0.354 |
| Native American | | 1.37 | 0.64 | 2.73 | 0.398 |
| Asian | | 2.48 | 0.49 | 9.38 | 0.216 |
| Pacific Islander | | 2.67 | 0.71 | 8.26 | 0.106 |
| African American, NOHO | | 1.16 | 0.93 | 1.45 | 0.205 |
| African American, OHO | | 1.16 | 0.79 | 1.68 | 0.447 |
| European American, NOHO | | 1.14 | 0.91 | 1.44 | 0.251 |
| European American, OHO | | 0.94 | 0.70 | 1.26 | 0.678 |
| **Education Level** | | **0.81** | **0.79** | **0.84** | **1.17E-45** |
| Household Income | | 0.95 | 0.92 | 0.99 | 0.007 |
| **MDD** | | **1.76** | **1.51** | **2.06** | **4.54E-13** |
| GAD | | 1.14 | 0.66 | 1.88 | 0.625 |
| **PTSD** | | **1.72** | **1.48** | **2.00** | **2.55E-12** |
| **OUD** | Severity | 0.99 | 0.77 | 1.26 | 0.920 |
|  | **Hazardous** | **1.73** | **1.41** | **2.13** | **2.05E-07** |
|  | Social Problems | 1.14 | 0.81 | 1.62 | 0.442 |
|  | Neglected Roles | 1.28 | 0.99 | 1.66 | 0.061 |
|  | Withdrawal | 1.35 | 0.90 | 2.03 | 0.154 |
|  | Tolerance | 1.09 | 0.81 | 1.47 | 0.589 |
|  | Larger Amounts | 0.71 | 0.54 | 0.95 | 0.019 |
|  | Attempts Quit | 0.62 | 0.45 | 0.86 | 0.005 |
|  | Time Spent | 0.80 | 0.57 | 1.12 | 0.182 |
|  | Physical Problems | 0.90 | 0.64 | 1.29 | 0.570 |
|  | Activities Given Up | 1.36 | 1.01 | 1.84 | 0.046 |
|  | Craving | 1.13 | 0.86 | 1.50 | 0.389 |

**Supplemental Table 9.** Association of antisocial personality disorder with severity and diagnostic criteria of tobacco use disorder (TUD). Associations surviving Bonferroni multiple testing correction (p < .002) are indicated in red. Covariates and phenotypes of interest are highlighted in orange and blue, respectively. NOHO: Not of Hispanic Origin; OHO: Of Hispanic Origin; MDD: major depressive disorder; GAD: generalized anxiety disorder; PTSD: posttraumatic stress disorder.

| **Variable** | | **OR** | **LCI** | **UCI** | ***p*** |
| --- | --- | --- | --- | --- | --- |
| Sex | | 1.00 | 0.87 | 1.14 | 0.947 |
| Age | | 1.00 | 0.99 | 1.00 | 0.471 |
| Native American | | 1.35 | 0.62 | 2.75 | 0.424 |
| Asian | | 2.74 | 0.51 | 11.01 | 0.188 |
| Pacific Islander | | 2.92 | 0.77 | 9.20 | 0.083 |
| African American, NOHO | | 1.12 | 0.90 | 1.41 | 0.314 |
| African American, OHO | | 1.04 | 0.71 | 1.51 | 0.850 |
| European American, NOHO | | 1.15 | 0.92 | 1.45 | 0.222 |
| European American, OHO | | 0.96 | 0.71 | 1.29 | 0.782 |
| **Education Level** | | **0.85** | **0.82** | **0.87** | **9.46E-28** |
| Household Income | | 0.97 | 0.94 | 1.01 | 0.141 |
| **MDD** | | **1.71** | **1.46** | **1.99** | **1.34E-11** |
| GAD | | 1.26 | 0.73 | 2.11 | 0.390 |
| **PTSD** | | **1.52** | **1.30** | **1.78** | **8.83E-08** |
| **TUD** | Severity | 1.05 | 0.87 | 1.28 | 0.598 |
|  | **Hazardous** | **1.88** | **1.62** | **2.18** | **1.14E-16** |
|  | Withdrawal | 1.13 | 0.94 | 1.36 | 0.183 |
|  | Tolerance | 1.16 | 0.99 | 1.37 | 0.067 |
|  | Larger Amounts | 1.03 | 0.83 | 1.26 | 0.812 |
|  | Attempts Quit | 1.02 | 0.83 | 1.24 | 0.865 |
|  | Time Spent | 0.98 | 0.83 | 1.15 | 0.777 |
|  | **Physical Problems** | **1.29** | **1.12** | **1.49** | **6.16E-04** |
|  | Activities Given Up | 1.14 | 0.96 | 1.34 | 0.126 |
|  | Craving | 1.02 | 0.87 | 1.19 | 0.821 |

**Supplemental Table 10.** Association of antisocial personality disorder with severity and diagnostic criteria of alcohol use disorder (AUD), cannabis use disorder (CanUD), cocaine use disorder (CocUD), opioid use disorder (OUD), and tobacco use disorder (TUD). The analysis was limited to diagnostic criteria that survived Bonferroni multiple testing correction in the SUD-specific analyses (Supplemental Tables 5, 6, 7, 8, and 9). Associations surviving Bonferroni multiple testing correction (p < 1.6E-03) are indicated in red. Covariates and phenotypes of interest are highlighted in orange and blue, respectively. NOHO: Not of Hispanic Origin; OHO: Of Hispanic Origin; MDD: major depressive disorder; GAD: generalized anxiety disorder; PTSD: posttraumatic stress disorder.

| **Variable** | | **OR** | **LCI** | **UCI** | ***p*** |
| --- | --- | --- | --- | --- | --- |
| **Sex** | | **1.33** | **1.15** | **1.54** | **1.15E-04** |
| Age | | 1.00 | 1.00 | 1.01 | 0.622 |
| Native American | | 1.46 | 0.66 | 3.01 | 0.321 |
| Asian | | 3.61 | 0.66 | 15.21 | 0.102 |
| Pacific Islander | | 3.04 | 0.78 | 9.91 | 0.080 |
| African American, NOHO | | 1.14 | 0.91 | 1.45 | 0.259 |
| African American, OHO | | 1.12 | 0.75 | 1.65 | 0.576 |
| European American, NOHO | | 1.01 | 0.80 | 1.29 | 0.907 |
| European American, OHO | | 0.98 | 0.72 | 1.33 | 0.887 |
| **Education Level** | | **0.85** | **0.82** | **0.87** | **1.10E-26** |
| Household Income | | 1.00 | 0.96 | 1.03 | 0.830 |
| **MDD** | | **1.63** | **1.39** | **1.91** | **1.61E-09** |
| GAD | | 1.21 | 0.69 | 2.06 | 0.480 |
| **PTSD** | | **1.32** | **1.12** | **1.54** | **6.54E-04** |
| AUD | Severity | 1.10 | 0.98 | 1.23 | 0.090 |
|  | Hazardous | 1.34 | 1.11 | 1.61 | 0.002 |
|  | Social Problems | 1.36 | 1.10 | 1.70 | 0.005 |
|  | Attempts Quit | 0.83 | 0.70 | 0.98 | 0.027 |
| **CanUD** | **Severity** | **1.17** | **1.08** | **1.26** | **3.66E-05** |
|  | Hazardous | 1.10 | 0.95 | 1.29 | 0.207 |
|  | Social Problems | 1.20 | 1.03 | 1.40 | 0.022 |
|  | Neglected Roles | 1.22 | 1.04 | 1.44 | 0.017 |
| **CocUD** | Severity | 1.06 | 0.95 | 1.18 | 0.276 |
|  | Hazardous | 1.23 | 1.05 | 1.45 | 0.012 |
|  | Neglected Roles | 1.19 | 1.00 | 1.42 | 0.045 |
|  | **Attempts Quit** | **0.64** | **0.50** | **0.83** | **8.00E-04** |
| OUD | Severity | 0.98 | 0.91 | 1.05 | 0.583 |
|  | Hazardous | 1.12 | 0.90 | 1.39 | 0.308 |
| **TUD** | Severity | 1.05 | 0.97 | 1.13 | 0.239 |
|  | **Hazardous** | **1.54** | **1.33** | **1.79** | **7.38E-09** |
|  | Physical Problems | 1.18 | 1.02 | 1.36 | 0.022 |

**Supplemental Table 11.** Association of antisocial personality disorder with severity and diagnostic criteria of alcohol use disorder (AUD), cannabis use disorder (CanUD), cocaine use disorder (CocUD), opioid use disorder (OUD), and tobacco use disorder (TUD). The model included diagnostic criteria that survived Bonferroni multiple testing correction in the SUD-specific analyses (Supplemental Tables 5, 6, 7, 8, and 9). Covariates, sex interaction terms, and phenotypes of interest are highlighted in orange, green, and blue, respectively. Associations among the phenotypes of interest maintaining nominal significance (p < 0.05) after the inclusion of sex interaction terms are indicated in red. NOHO: Not of Hispanic Origin; OHO: Of Hispanic Origin; MDD: major depressive disorder; GAD: generalized anxiety disorder; PTSD: posttraumatic stress disorder.

| **Variable** | | **OR** | **LCI** | **UCI** | | ***p*** | |
| --- | --- | --- | --- | --- | --- | --- | --- |
| Sex | | 1.14 | 0.86 | 1.51 | | 0.362 | |
| Age | | 1.00 | 1.00 | 1.01 | | 0.628 | |
| Native American | | 1.45 | 0.66 | 2.98 | | 0.334 | |
| Asian | | 3.64 | 0.66 | 15.33 | | 0.101 | |
| Pacific Islander | | 3.07 | 0.78 | 10.07 | | 0.079 | |
| African American, NOHO | | 1.14 | 0.91 | 1.45 | | 0.255 | |
| African American, OHO | | 1.12 | 0.75 | 1.65 | | 0.566 | |
| European American, NOHO | | 1.01 | 0.80 | 1.29 | | 0.910 | |
| European American, OHO | | 0.98 | 0.72 | 1.33 | | 0.900 | |
| Education Level | | 0.85 | 0.82 | 0.87 | | 1.02E-26 | |
| Household Income | | 1.00 | 0.96 | 1.03 | | 0.852 | |
| MDD | | 1.63 | 1.39 | 1.91 | | 1.32E-09 | |
| GAD | | 1.23 | 0.70 | 2.09 | | 0.452 | |
| PTSD | | 1.31 | 1.12 | 1.53 | | 7.72E-04 | |
| AUD | Severity | 1.10 | 0.98 | | 1.23 | | 0.097 |
|  | Hazardous | 1.33 | 1.11 | | 1.60 | | 0.002 |
|  | Social Problems | 1.37 | 1.10 | | 1.70 | | 0.005 |
|  | Attempts Quit | 0.83 | 0.70 | | 0.98 | | 0.027 |
| CanUD | Hazardous | 1.11 | 0.95 | | 1.30 | | 0.187 |
|  | Social Problems | 1.20 | 1.03 | | 1.41 | | 0.020 |
|  | Neglected Roles | 1.23 | 1.04 | | 1.45 | | 0.015 |
| CocUD | Severity | 1.06 | 0.95 | | 1.18 | | 0.282 |
|  | Hazardous | 1.23 | 1.05 | | 1.45 | | 0.013 |
|  | Neglected Roles | 1.19 | 1.01 | | 1.42 | | 0.044 |
| OUD | Severity | 0.98 | 0.91 | | 1.05 | | 0.564 |
|  | Hazardous | 1.12 | 0.90 | | 1.40 | | 0.298 |
| TUD | Severity | 1.05 | 0.97 | | 1.13 | | 0.240 |
|  | Physical Problems | 1.18 | 1.02 | | 1.36 | | 0.024 |
| Sex 🞨 CanUD Severity | | 1.05 | 0.94 | 1.17 | | 0.423 | |
| Sex 🞨 CocUD Attempts Quit | | 1.11 | 0.81 | 1.51 | | 0.515 | |
| Sex 🞨 TUD Hazardous | | 1.06 | 0.79 | 1.40 | | 0.710 | |
| CanUD Severity | | 1.10 | 0.94 | 1.29 | | 0.232 | |
| **CocUD Attempts Quit** | | **0.57** | **0.36** | **0.89** | | **0.013** | |
| TUD Hazardous | | 1.44 | 0.98 | 2.12 | | 0.061 | |
